# Beyond Healthcare Utilization: A Whole-Child, Multi-Sector Approach to Pediatric Risk Stratification

**DOI:** 10.64898/2026.09.08.26361653

**Authors:** Anna F. Zolotor, Alex Gertner, Richard J. Chung, Josie Hatley, Chelsea Swanson, Kori B. Flower, Michael J. Steiner, John Bainbridge, Chavanne Lamb, Eleanor Wertman, Rushina Cholera

## Abstract

**Objectives:** The North Carolina Integrated Care for Kids (NC InCK) pediatric demonstration model created a whole-child risk stratification approach to identify children for enhanced care management. Unlike traditional approaches that rely primarily on healthcare data, NC InCK integrates health, education, social, and juvenile justice data. We described NC InCK’s risk stratification approach and compared its output to Medicaid managed care organization (MCO)-assigned risk levels, with the hypothesis that these approaches would identify distinct populations.

**Methods:** This was a cross-sectional study including NC Medicaid participants aged 0-20 years residing in a five-county region in central NC in October 2022. NC InCK’s risk stratification algorithm was used to assign participants to Service Integration Levels (SILs) 1, 2, or 3, reflecting increasing anticipated need for integrated cross-sector support. Descriptive analyses compared participant baseline characteristics by NC InCK and Medicaid MCO risk level.

**Results:** Of 99,564 children, 90,151 (90.5%) were assigned to SIL 1, 5,482 (5.5%) to SIL 2, and 3,931 (4%) to SIL 3. About 24% and 20% of children assigned to SILs 2 or 3 by NC InCK were identified as low risk by their Medicaid MCO.

**Conclusions:** NC InCK’s risk stratification approach assigns children to SILs 2 and 3 using health, education, social, and juvenile justice data. These populations differ from those identified by Medicaid MCO risk stratification, suggesting that inclusion of non-clinical variables adds unique value. This approach is a step forward in demonstrating the feasibility of methods that proactively identify populations for preventive support by including upstream risk factors.

## Introduction

Risk stratification is used by health systems and payers to identify populations at risk of poor health outcomes and proactively target care delivery and tailored population health interventions.^1^ While risk stratification is used for both adult and pediatric populations, most approaches were developed for adults and emphasize chronic conditions, healthcare utilization and healthcare spending.^1^ Children represent a distinct population with generally lower healthcare utilization and expenditures than adults; high costs are concentrated in a small proportion of medically complex children.^2–4^ While family well-being and health-related social needs (HRSNs) in childhood predict lifelong health and well-being,^5^ these factors are rarely incorporated into risk stratification. Traditional stratification approaches might under-identify children whose unmet HRSNs or limited access to care place them at risk for adverse outcomes despite low healthcare utilization.^6^

We describe a novel whole-child risk stratification approach developed as a component of the North Carolina Integrated Care for Kids (NC InCK) model. NC InCK is one of seven InCK local service delivery and state payment models for Medicaid-enrolled children funded by the Centers for Medicare and Medicaid Innovation (CMMI).^7^ CMMI designed the InCK models to improve child health and reduce avoidable hospitalization and out-of-home placements (OOHP) through coordinated, cross-sector approaches focused on prevention and early intervention. To support a comprehensive assessment of children’s needs and early identification of those who may benefit from proactive support, NC InCK developed a risk stratification approach that integrates healthcare, education, guardian, HRSN, and juvenile justice system data. This approach was designed to complement the risk stratification approaches used by MCOs, which are typically driven primarily by clinical indicators and healthcare utilization data.^8,9^ Children are assigned to Service Integration Level (SIL) 1, 2, or 3, where higher SIL indicates greater potential to benefit from integrated health and social supports. Children assigned to SILs 2 and 3 are prioritized for outreach to offer NC InCK’s multidimensional longitudinal care management program, which is designed to address clinical, social and family needs.^10^

This paper describes the NC InCK risk stratification approach, characterizes children based on Service Integration Level (SIL) assignment, and compares the populations identified by NC InCK with those identified by Medicaid managed care organizations (MCOs). By examining differences between these approaches, we assess whether a whole-child model identifies children who may be overlooked by traditional utilization-based methods. This work has implications for Medicaid programs and health systems seeking to leverage integrated administrative cross-sector data to support proactive, whole-child care.

## Methods

### Design Context

NC InCK’s risk stratification approach (Exhibit 1) was developed through an iterative consensus-building process by an expert panel including medical and behavioral healthcare providers, representatives from cross-sector child-serving agencies (e.g. social services, child welfare), NC Medicaid and child health subject matter experts. Reflecting CMMI InCK priorities, there was a focus on identifying children with current OOHP (including foster care or juvenile justice detention) and those at risk of preventable OOHP based on complex health and social needs. As children with medical complexity are already prioritized through MCO risk stratification approaches, NC InCK’s method was designed to complement existing strategies by identifying children whose combined health and social risks may not be captured by traditional models. Therefore, the approach was designed to identify children with *concurrent health and social risk factors* and did not assign children to SIL 2 or 3 based on health risks alone.

SIL assignments are updated monthly and children in SIL 2 or 3 are prioritized for outreach to offer care management. To align with operational care management capacity, the algorithm was designed to assign fewer than 15% of the population to SIL 2 or 3. Beginning in October 2022, NC InCK’s risk stratification method was applied to all Medicaid-enrolled children ages 0 to 20 years residing in NC InCK’s five-county service region in central NC (approximately 100,000 children). Analyses presented here reflect the baseline SIL assignments made in October 2022.

### Data Sources

The risk stratification method incorporates NC Medicaid data from the NC Department of Health and Human Services (DHHS), education data from the NC Department of Public Instruction (DPI), and juvenile justice information from the NC Department of Public Safety (DPS). The NC Government Data Analytics Center (GDAC) receives and links data across agencies, applies the risk stratification approach, and transmits risk levels to NC InCK. Data sharing and use were governed by formal interagency agreements among participating state agencies and were conducted in accordance with NC InCK program requirements.

### SIL Assignment

NC InCK’s risk stratification approach uses 34 binary indicators grouped into seven domains: Current OOHP, OOHP Risk, Healthcare Designation, Healthcare Utilization, Socioeconomic, Education, and Guardian (Exhibit 2). For each domain, a positive value was assigned when any indicator in that domain was present. For example, more than 45 school absences in the past year is one of four indicators within the education domain. A child with more than 45 absences in the past year would be classified as positive for the education domain, regardless of the status of the remaining education indicators.

SIL assignment follows a five-step sequence (Exhibit 1):

1. Children with current OOHP are assigned to SIL 3
2. Among remaining children, those without either a positive health designation or healthcare utilization indicator are assigned to SIL 1
3. Children meeting criteria for risk of OOHP are assigned to SIL 3
4. Among remaining children, those with positive indicators in *all three* social-contextual domains (Socioeconomic, Education, and Guardian) are assigned to SIL 3
5. Remaining children with positive indicators in *any two* social-contextual domains are assigned to SIL 2; all others are assigned to SIL 1

### Measures and Statistical Analysis

We describe population characteristics overall and by NC InCK SIL, including gender, age, race, ethnicity, Social Deprivation Index (SDI) score, Medicaid costs, and Medicaid MCO risk levels. SDI score, a validated area-level measure of deprivation, was determined based on the census tracts where children resided.^11^ Medicaid costs were calculated using total paid claims over the prior 12-month period. Medicaid MCO risk levels (low, medium, or high) were assigned by each child’s MCO. While NC Medicaid requires all MCOs to risk stratify beneficiaries using data elements including claims data, clinical results, utilization and demographics, plans are not required to share details of their methods.^12,13^ However, payers typically use methods such as the Johns Hopkins Adjusted Clinical Groups System, which relies on healthcare utilization, cost, diagnoses, and some demographic characteristics.^8,9^

To compare the populations identified by NC InCK and MCO risk stratification, we examined the distribution and characteristics of children across Medicaid risk levels. Because children enrolled in Medicaid Direct (fee-for-service Medicaid) are eligible for NC InCk but are not included in MCO risk stratification, they were analyzed as a separate group. This population includes children in foster care, medically fragile individuals and other groups with specialized healthcare needs who are excluded from MCO enrollment.^14^

Analyses were conducted using SAS v 9.5. The project was deemed exempt by the Institutional Review Boards at Duke University and the University of North Carolina at Chapel Hill.

## Results

### Population Characteristics

In October 2022, 99,564 children were eligible for NC InCK based on age, county, and enrollment in Medicaid or the Children’s Health Insurance Program (CHIP). Of these children, 50.4% (N=50,203) were male. The largest age groups were 5-11 (34.5%; N=34,332) and 12-17 years old (29.1%; N=29,014) (Exhibit 2). Most children were white (51.1%; N=50,908) followed by Black or African American (41.7%; N=41,480). Approximately one-third of children (30.3%; N=30,153) were of Hispanic or Latine ethnicity. The median SDI score was 73 (IQR: 44), where 0 indicates the lowest deprivation and 100 indicates the highest deprivation. The median annual cost paid by Medicaid was $790.09 (IQR: $1,438.68).

### NC InCK SIL Assignment

Of the 99,564 children, 90.5% (N=90,151) were assigned to SIL 1, 5.5% (N= 5,482) to SIL 2 and 3.9% (N=3,931) to SIL 3. Of the 3,931 children in SIL 3, 20.4% (N=802) qualified based on current OOHP, 59.9% (N=2,354) based on a positive health-related category and OOHP risk, and 19.7% (N=775) based on a positive health-related category *and* all three social-contextual categories. 5,482 children were assigned to SIL 2 based on having at least one positive health-related category and two positive social-contextual categories.

Among the 5,482 children assigned to SIL 2, the distribution of positive social-contextual category combinations varied. In particular, 68.6% (N=3,763) of children had concurrent positive Guardian and SES categories, 24.7% (N=1,352) had positive SES and Education categories, and 6.7% (N=367) had positive Education and Guardian categories.

### Characteristics by NC InCK Risk Level

On average, children assigned to SIL 3 were 0.76 and 1.65 years older than those in SIL 2 and SIL 3, respectively. Over half of children in SIL 2(54.9%; N=3,011) and SIL 3 (53.4%; N=2,097) were Black. By contrast, Hispanic/Latine children were a smaller proportion of SIL 2(13.9%; N=764) and SIL 3 (10.2%; N=401) compared to the overall population. The median SDI score was 72 for SIL 1, 86 for SIL 2, and 73 for SIL 3. Mean annual paid Medicaid cost was highest for SIL 3 ($12,784.24), followed by SIL 2($4,238.81) and SIL 1($724.04), while the median cost was similar for SIL 2 and 3 ($2,199.79 [IQR: $3,288.1] and $2,520.88 [IQR: $6,547.34], respectively).

### Medicaid MCO Risk Levels

Risk stratification by the Medicaid MCOs assigned 41.0% (N=40,786) of children to the low risk level, 26.3% (N=26,169) to medium risk, and 5.7% (N=5,686) to high risk (Supplement). A further 9.7% (N=9,698) of children eligible for NC InCK had Medicaid Direct. For analyses examining concordance between MCO and NC InCK SIL assignments, children enrolled in Medicaid Direct were grouped with children assigned to the Medicaid high risk category. The remaining 17.3% (N=17,225) had missing MCO-assigned risk levels, an allowable entry when plans do not yet have enough information to make a risk level designation.

### NC InCK SILs compared to Medicaid MCO Risk Levels

NC InCK SILs and Medicaid MCO risk level assignments were concordant for 42.9% (N=38,690) of NC InCK SIL 1 children, 30% (N=1,642) of SIL 2 children, and 65.7% (N=2,584) of SIL 3 children (grouping Medicaid Direct with MCO high risk) (Exhibit 4). Concordance was lower among children assigned to SIL 2 or SIL 3 by NC InCK. Of the 90,151 children assigned to SIL 1, 26.8% (N= 24,192) were assigned to the MCO medium risk level, 5.2% (N=4,665) to the Medicaid high risk level, and 7.1% (N=6,404) to Medicaid Direct. Of the 5,482 children assigned to SIL 2, 24.3% (N=1,334) were assigned to the Medicaid low risk level, 8.8% (N=481) to high, and 22.80% (N=1,250) to Medicaid Direct. Fewer children assigned to SIL 3 had missing MCO risk levels relative to SIL 1 (6.4% versus 18%). The proportion of children with Medicaid Direct increased with higher NC InCK SIL, with 52% of those assigned to SIL 3 having Medicaid Direct.

As with the NC InCK SILs, there was a higher proportion of Black children in the MCO high risk group (56.7%; N=3,225) compared with the low risk group (36.7%; N=14,983); the proportion of Black children was similar for the Medicaid medium risk (44.9%; N=11,746) and Medicaid Direct (44.5%; N=4,682) groups. In contrast to the NC InCK SILS, the proportions of Hispanic/Latine children were similar for low (29.7%; N=12,139), medium (35.6%; N=26,169), and high risk MCO groups (29.9%; N=1702); the proportion of Hispanic/Latine children was lower in the Medicaid Direct group (18.4%; N=1,783- ). While Medicaid costs were comparable between the MCO low risk group and NC InCK SIL 1 ($1,864.24 versus $1,750.50), costs in SILs 2 and 3 ($4,248.81 and $12,784.24, respectively) were higher than the MCO medium and high risk groups ($1,946.77 and $5,580.86, respectively).

## Discussion

Risk stratification is a key population health strategy used to identify patients at increased risk of adverse outcomes and guide targeted interventions. However, most approaches are reactive and oriented toward adults with frequent or costly healthcare utilization. We describe NC InCK’s whole-child risk stratification approach which integrates healthcare, educational, social and contextual data to identify children who may benefit from additional support. This approach assigned approximately 10% of the eligible pediatric population to SIL 2 or 3. Children in NC InCK SILs 2 and 3 differed in demographic characteristics from children in SIL 1; furthermore, NC InCK and Medicaid MCO algorithms elevated distinct groups of children.

The variation in demographic characteristics across NC InCK risk groups reflects disparities in health and wealth and can serve as a useful baseline for future pediatric risk stratification development. On average, children designated to SILs 2 and 3 were slightly older than the overall population, likely because risk indicators such as school absences do not appear until children have interacted with education systems. Children assigned to these groups were also more likely to be Black, reflecting the effects of structural racism, which leads to higher rates of material deprivation and poorer health.^15^ By comparison, algorithms that rely primarily on healthcare utilization data may fail to appropriately identify minoritized populations who face systemic barriers to accessing healthcare. For example, a large commercial risk stratification tool was shown to perpetuate racial bias by using healthcare costs as a proxy for health, because less money is spent on equally sick Black patients compared to White patients.^16^

Hispanic or Latine children were under-represented in NC InCK SILs 2 and 3, a finding which could reflect true differences in risk, differential engagement with systems, or both. In particular, this finding may in part reflect the Hispanic health paradox, wherein Latine populations have better overall health than other demographic groups despite low-income status.^17^ Alternatively, Latine individuals may be less likely to engage with healthcare, education, and other service systems incorporated into the NC InCK stratification approach due to language barriers, cultural differences, and discriminatory practices.^18,19^ Caregivers of Latine children enrolled in NC Medicaid have reported substantially lower trust in healthcare providers compared to caregivers of non-Latine children.^20^ Importantly, NC InCK’s stratification approach includes an indicator for the *absence* of preventive healthcare visits in children under age five as one method to identify children with barriers to healthcare. Pairing approaches that integrate cross-sector data with direct referrals from trusted community partners or clinicians may help identify and engage families who have limited interaction with public systems, although such approaches can also be susceptible to provider-level biases.^21,22^ Careful examination of unintentional bias is necessary for any risk stratification approach.^23^

The children prioritized by NC InCK differed substantially from those identified as high or medium risk by Medicaid MCOs, likely reflecting differences in the constructs measured by each approach. Traditional risk stratification models primarily use health/healthcare data to identify children at risk for high healthcare utilization or expenditures; NC InCK’s approach was designed to identify children with concurrent health and social risks who may not be elevated based on health-related data alone. Importantly, NC InCK SIL assignments do not replace or supersede Medicaid MCO risk classifications in practice. Children identified as high risk by their Medicaid MCO remain eligible for MCO care management and related services regardless of their NC InCK SIL assignment. These approaches can therefore be viewed as complementary.

Notably, children assigned to SILs 2 and 3 had higher average Medicaid expenditures than children assigned to the Medicaid MCO medium and high risk categories, despite the NC InCK algorithm requiring indication of health/healthcare *and* social-contextual risk for elevation. This finding should be interpreted cautiously but may suggest that approaches based on a combination of cross-sector social-contextual data and healthcare data can more effectively distinguish children with complex, multi-dimensional needs than approaches based primarily on healthcare utilization and claims data. Future work will examine whether NC InCK risk designations are more predictive of subsequent health and social outcomes and whether alternative sequencing or weighting of health and social indicators improves performance.

The NC InCK risk stratification approach identifies children who might otherwise be overlooked by traditional risk stratification methods largely because of the integration of multisector state-level data. NC InCK’s approach is intended to leverage these additional data sources to identify children who may be at elevated risk for future adverse outcomes *before* those risks manifest as high healthcare utilization, hospitalization, or out-of-home placement. While data integration efforts can have high upfront costs, effective early intervention has potential to yield public savings in foster care, juvenile justice services, and treatment of chronic diseases.^24–27^

### Limitations

Our study focused on a Medicaid-enrolled pediatric population residing in central NC. As such, the distribution of risk profiles is not necessarily generalizable to other populations. Our population had few Native Americans, Alaskans, or Pacific Islanders, so we could not describe how the approach functioned for these groups. We did not have access to details of the stratification methods used by Medicaid MCOs, limiting our ability to draw conclusions based on comparisons with the NC InCK approach.

## Conclusion

NC InCK’s whole-child risk stratification approach reflects a holistic view of pediatric risk that incorporates health, social, family, and educational factors rather than relying primarily on healthcare utilization and cost data. This approach identifies a distinct population of children compared to a traditional risk stratification approach, suggesting that cross-sector whole-child frameworks may help identify children who can benefit from proactive intervention to prevent future adverse health outcomes. Future work should evaluate whether incorporating cross-sector data improves prediction of subsequent outcomes, supporting more effective and efficient population health management strategies for children and families.

**Figure 1:**
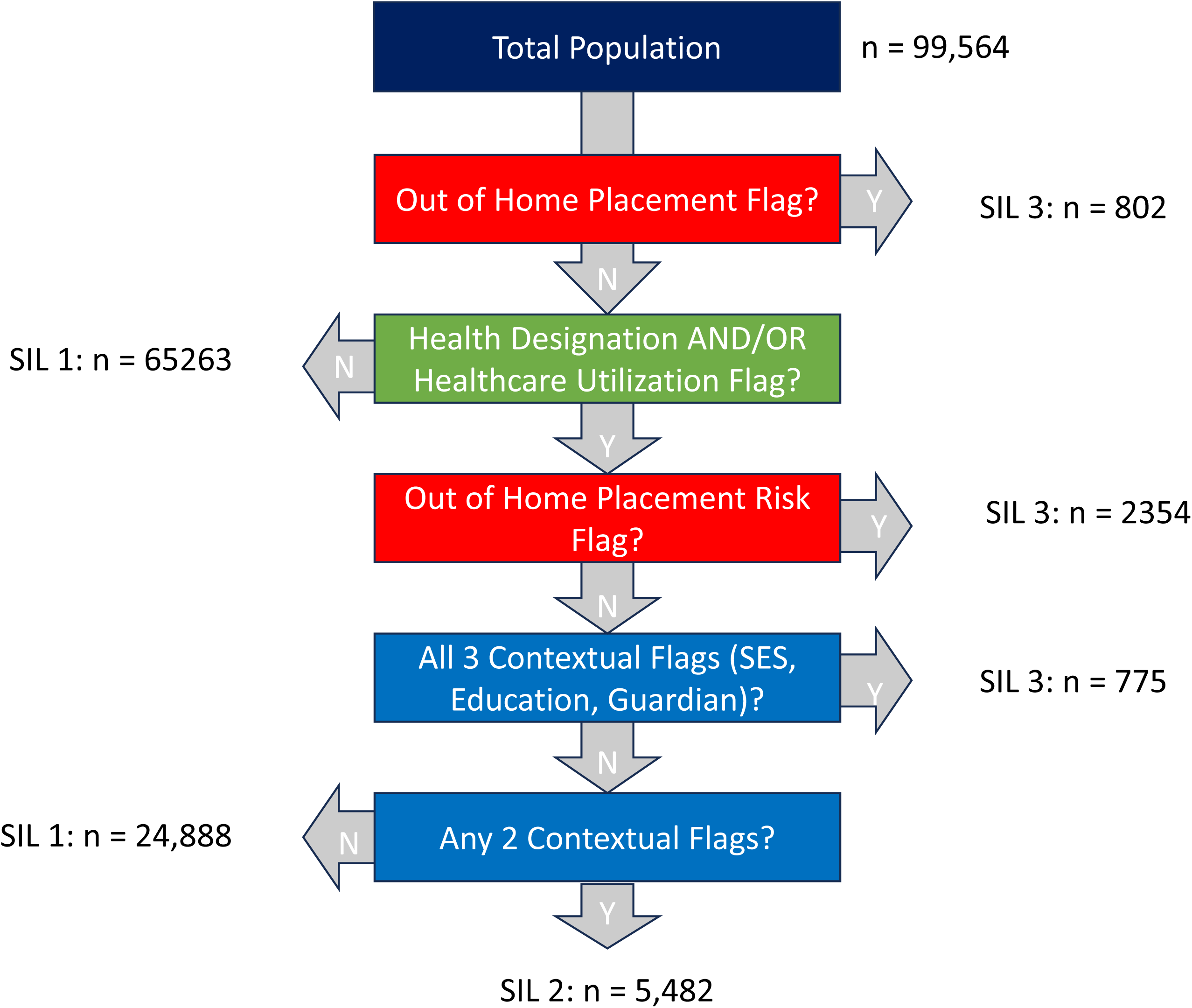
Step-by-step assignment of Service Integration Levels in the NC InCK risk stratification approach.

**Figure 2:**
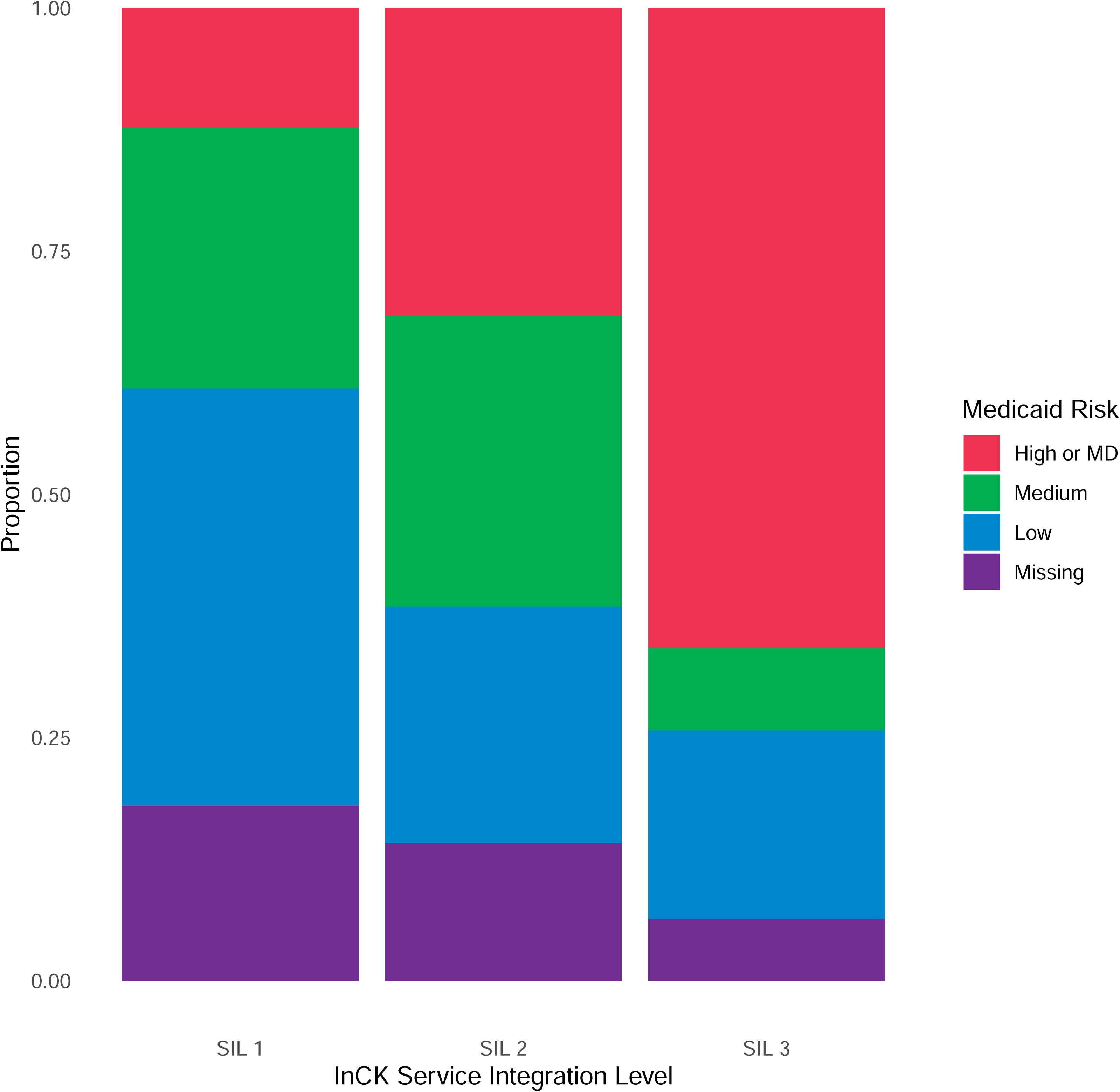
Medicaid Managed Care Organization risk levels for children in each InCK Service Integration Level. Footnote: Children with MD (Medicaid Direct) are combined with children assigned to Medicaid high risk level for the purpose of this visualization.

**Table 1:** Variables in the NC InCK risk stratification algorithm by Category.

| Variables in each Category | Definition of “yes” for this element |
| --- | --- |
| **Current Out of Home Placement (Step 1)** |  |
| Juvenile justice residential placement (current) | Indicator of placement in a detention facility, a youth development center, or other JJ residential programming currently or within the past 12 months |
| Foster care plan enrollee (current) | Currently enrolled in the Medicaid Direct foster care plan |
| **Out of Home Placement Risk (Step 3)** |  |
| Prior foster care plan enrollee | Indicates previous enrollment in foster care plan which has since ended in the last 12-month period. |
| Adoption assistance plan enrollee | Indicates new entry into the adoption assistance plan in the last 12 months. |
| Juvenile justice involvement | Any juvenile justice involvements, including Residential placement or other higher-severity, |
| Two or more hospitalizations in past year | Two or more hospitalizations within one year during the period from 14-2 months prior to run date. |
| 15 or more inpatient days within past year | 15 or more inpatient days during the period from 14-2 months prior to run date. |
| Residential treatment within past year | One or more days in residential treatment during the period from 14-2 months prior to run date. |
| Skilled nursing facility treatment within past year | One or more days in skilled nursing facility during the period from 14-2 months prior to run date. |
| Psychiatric inpatient admission within past year | One or more days in psychiatric inpatient admission during the period from 14-2 months prior to run date. |
| **Health Designation (Step 2)** |  |
| Care Management for At-risk Children (CMARC) enrollee | Enrolled in CMARC (a Medicaid care management program for at-risk children ages 0-5) at any time during prior 12 months. |
| Community Alternatives Program for Children (CAP/C) enrollee | Enrolled in CAP/C (a program that provides an alternative to institutionalization for medically fragile Medicaid beneficiaries) at any time during prior 12 months. |
| Pediatric Medical Complexity Algorithm (PMCA) 3.0 level 2 or greater | Level 2 or 3 on most recent PMCA score (indicates complex chronic disease or non-complex chronic disease) at time of stratification. Updated in January and July of every year using previous 3 years of data. |
| Tailored Plan Eligible | Currently eligible for Tailored Plan (a managed care plan for those with behavioral health complexity). |
| **Healthcare Utilization (Step 2)** |  |
| Antipsychotic prescription within past year | One or more antipsychotic prescriptions dispensed to member during the period from 14-2 months prior to run date. |
| In-home mental health services within past year | Indicator of any receipt of in-home mental health services during the period from 14-2 months prior to run date. |
| Mobile crisis response use within past year | One or more claims for mobile crisis response utilization during the period from 14-2 months prior to run date. |
| Three or more emergency room visits within past year | Three or more ER visits during period 14-2 months prior to run date. |
| Medicaid paid cost > \$2735 within past year | Total paid cost of greater than \$2735 on behalf of member during the period from 15-3 months prior to run date. |
| Less than 5 years old without any claims despite continuous enrollment for 2 years | Indicator for children 4 or younger who have been continuously enrolled in Medicaid during the period from 26-2 months prior to the date of run but has not had any claims during that time. |
| **Socioeconomic (Step 4, 5)** |  |
| Temporary Assistance for Needy Families (TANF) eligible | Currently eligible for TANF (cash assistance program for low-income families with children; eligibility is an indicator of substantial hardship). |
| Social Deprivation Index, high score | SDI (a measure of community deprivation based on census tract data<sup>28</sup>) score is 90 or above on a scale of 0-100 based on current patient address. |

| **Education (Step 4, 5)** |  |
| --- | --- |
| Chronic school absences within last academic year | Absent more than 45 days within most recent year of included schools data (threshold specific to 2019-2020 schools data). |
| Extended suspensions within past academic year | Four or more days of in-school-suspension, 9 or more days of short-term suspension, or any days of long-term suspension within most recent year of included schools data (threshold specific to 2019-2020 schools data). |
| Expulsion within past academic year | Child was expelled during most recent year of included schools data. |
| Early intervention Infant-Toddler Program within past year | Enrolled in North Carolina Infant-Toddler Program (provides support for families and children from birth to three who have special needs) or associated services billed during 12 months prior to run date. |
| **Guardian (Step 4, 5)** |  |
| Guardian tailored plan eligible in past 2 years | Guardian eligible for Tailored Plan during the period from 26-2 months prior to run date. |
| Guardian psychiatric admission in past 2 years | Guardian has 1 or more behavioral health inpatient admissions during period from 2-14 months prior to run date. |
| Guardian Medicaid eligible due to disability in past 2 years | Guardian eligible for Medicaid due to disability during the period from 26-2 months prior to run date. |
| Guardian substance use during pregnancy in past 2 years | Maternal perinatal depression indicated during the period from 26-2 months prior to run date. |
| Guardian perinatal depression in past 2 years | Maternal perinatal depression indicated during the period from 26-2 months prior to run date. |

**Table 2:**
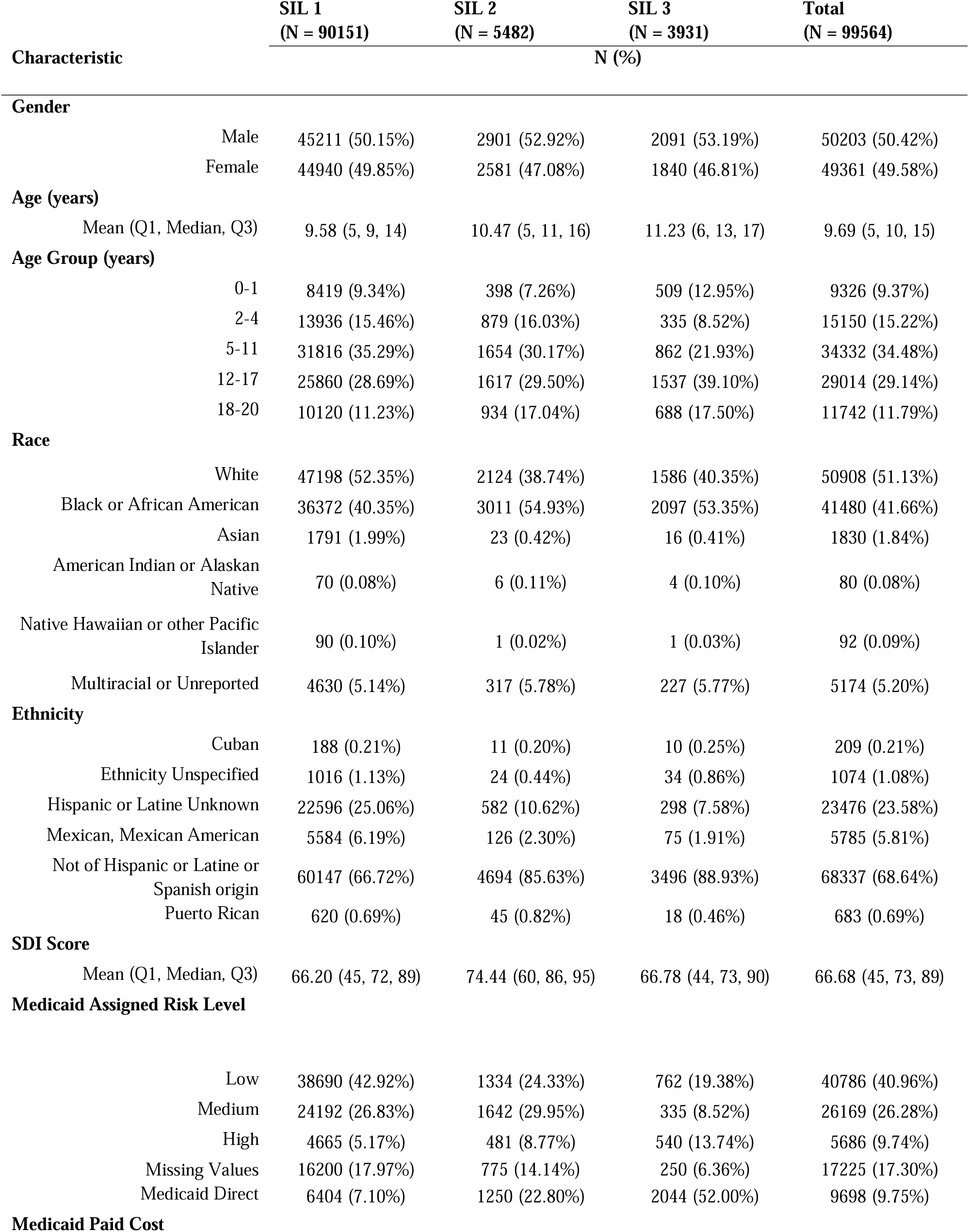
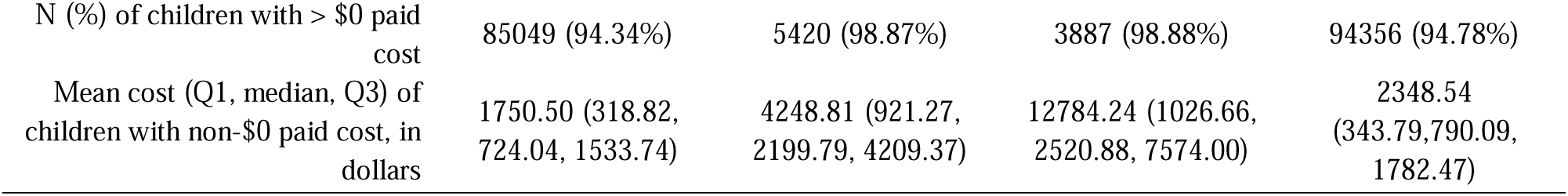
Characteristics of children by InCK Service Integration Level.

## Data Availability

The data used in the present study are not not publicly available.

## Acknowledgements

The authors acknowledge Sarah Allin (previously NC InCK), Charlene Wong (Duke Department of Pediatrics), and Adam Zolotor (University of North Carolina Department of Family Medicine) for their contributions in designing the NC InCK risk stratification algorithm.

