## Supplemental Table 1 for "Beyond Healthcare Utilization: A Whole-Child, Multi-Sector Approach to Pediatric Risk Stratification"

Supplement: Population characteristics, stratified by Medicaid Risk Level

|  | **Low Risk**  **(N = 40786)** | **Medium Risk**  **(N = 26169)** | **High Risk**  **(N = 5686)** | | **Medicaid Direct**  **(N = 9698)*** | **Risk level missing (N =17225)** | **Total**  **(N = 99564)** |
| --- | --- | --- | --- | --- | --- | --- | --- |
| **Characteristic** | | | | **N (%)** | | | |
| **Gender** |  |  |  | |  |  |  |
| Male | 20246 (49.64%) | 12951 (49.49%) | 2781 (48.89%) | | 5670 (58.47%) | 8556 (49.67%) | 50203 (50.42%) |
| Female | 20540 (50.36%) | 13218 (50.51%) | 2906 (51.11%) | | 4028 (41.53%) | 8669 (50.33%) | 49361 (49.58%) |
| **Age (years)** | | | | | | | |
| Mean (Q1, Median, Q3) | 9.45 (4, 9, 14) | 9.54 (5, 9, 14) | 9.58 (4, 9, 15) | | 12.41 (9, 13, 17) | 9.02 (4, 9, 14) | 9.69 (5, 10, 15) |
| **Age Group (years)** | | | | | | | |
| 0-1 | 3961 (9.71%) | 2191 (8.37%) | 745 (13.10%) | | 169 (1.74%) | 2260 (13.12%) | 9326 (9.37%) |
| 2-4 | 6666 (16.34%) | 4270 (16.32%) | 866 (15.23%) | | 656 (6.76%) | 2692 (15.63%) | 15150 (15.22%) |
| 5-11 | 14282 (35.02%) | 9412 (35.97%) | 1694 (29.79%) | | 3077 (31.73%) | 5867 (34.06%) | 34332 (34.06%) |
| 12-17 | 11274 (27.64%) | 7591 (29.01%) | 1611 (28.33%) | | 3909 (40.31%) | 4629 (26.87%) | 29014 (29.14%) |
| 18-20 | 4603 (11.29%) | 2705 (10.34%) | 770 (13.54%) | | 1887 (19.46%) | 1777 (10.32%) | 11742 (11.79%) |
| **Race** | | | | | | | |
| White | 22829 (55.97%) | 12703 (48.54%) | 2110 (37.11%) | | 4682 (48.28%) | 8584 (49.83%) | 50908 (51.13%) |
| Black or African American | 14983 (36.74%) | 11746 (44.89%) | 3225 (56.72%) | | 4314 (44.48%) | 7212 (41.87%) | 41480 (41.66%) |
| Asian | 919 (2.25%) | 357 (1.36%) | 25 (0.44%) | | 135 (1.39%) | 394 (2.29%) | 1830 (1.84%) |
| American Indian or Alaskan Native | 30 (0.07%) | 15 (0.06%) | 4 (0.07%) | | 23 (0.24%) | 8 (0.05%) | 80 (0.08%) |
| Native Hawaiian or other Pacific Islander | 34 (0.08%) | 8 (0.03%) | 0 (0.00%) | | 14 (0.14%) | 36 (0.21%) | 92 (0.09%) |
| Multiracial or Unreported | 1991 (4.88%) | 1340 (5.12%) | 322 (5.66%) | | 530 (5.47%) | 991 (5.75%) | 5174 (5.20%) |
| **Ethnicity** | | | | | | | |
| Cuban | 91 (0.22%) | 46 (0.18%) | 10 (0.18%) | | 24 (0.25%) | 38 (0.22%) | 209 (0.21%) |
| Ethnicity Unspecified | 465 (1.14%) | 179 (0.68%) | 41 (0.72%) | | 176 (1.81%) | 213 (1.24%) | 1074 (1.08%) |
| Hispanic or Latine Unknown | 9258 (22.70%) | 7332 (28.02%) | 1303 (22.92%) | | 1373 (14.16%) | 4210 (24.44%) | 23476 (23.58%) |
| Mexican, Mexican American | 2542 (6.23%) | 1739 (6.65%) | 348 (6.12%) | | 318 (3.28%) | 838 (4.87%) | 5785 (5.81%) |
| Not of Hispanic or Latine or Spanish origin | 28182 (69.10%) | 16675 (63.72%) | 3943 (69.35%) | | 7739 (79.80%) | 11798 (68.49%) | 68337 (68.64%) |
| Puerto Rican | 248 (0.61%) | 198 (0.76%) | 41 (0.72%) | | 68 (0.70%) | 128 (0.74%) | 683 (0.69%) |
| **SDI Score** | | | | | | | |
| Mean (Q1, Median, Q3 | 65.62 (44, 72, 87) | 68.38 (49, 75, 90) | 74.67 (62, 81, 93) | | 63.13 (40, 67, 87) | 65.91 (44, 72, 89) | 66.68 (45, 73, 89) |
| **InCK SIL** | | | | | | | |
| SIL 1 | 38690 (94.86%) | 24192 (92.45%) | 4665 (82.04%) | | 6404 (66.03%) | 16200 (94.05%) | 90151 (90.55%) |
| SIL 2 | 1334 (3.27%) | 1642 (6.27%) | 481 (8.46%) | | 1250 (12.89%) | 775 (4.50%) | 5482 (5.51%) |
| SIL 3 | 762 (1.87%) | 335 (1.28%) | 540 (9.50%) | | 2044 (21.08%) | 250 (1.45%) | 3931 (3.95%) |
| **Medicaid paid cost ($)** | | | | | | | |
| N (%) of children with $0 paid cost | 38950 (95.50%) | 25754 (98.41%) | 5552 (97.64%) | | 8659 (89.29%) | 15441 (89.64%) | 94356 (94.77%) |
| Mean (Q1, median, Q3) of children with non-$0 paid cost | $1864.24 (318.84, 732.75, 1592.28) | $1946.77 (328.68, 746.69, 1583.22) | $5580.86 (529.13, 1187.03, 3014.21) | | $4223.78 (429.85, 1202.83, 3028.74) | $2026.51(352.78, 770.04, 1722.33) | 2348.54  (343.79,790.09, 1782.47) |

*Approximately 10% of Medicaid Direct children had assigned Medicaid risk levels; however, these children are included in the Medicaid Direct column rather than in the columns associated with their assigned risk levels.
